# Deep immune profiling uncovers novel associations with clinical phenotypes of Multisystem Inflammatory Syndrome in Children (MIS-C)

**DOI:** 10.1101/2022.08.31.22279265

**Authors:** Christopher Redmond, Moses M. Kitakule, Aran Son, McKella Sylvester, Keith Sacco, Ottavia Delmonte, Francesco Licciardi, Riccardo Castagnoli, Cecilia Poli, Yasmin Espinoza, Camila Astudillo, Sarah E. Weber, Gina A. Montealegre Sanchez, Karyl Barron, Mary Magliocco, Kerry Dobbs, Yu Zhang, Helen Matthews, Cihan Oguz, Helen C. Su, Luigi D. Notarangelo, Pamela A. Frischmeyer-Guerrerio, Daniella M. Schwartz

**Affiliations:** Vasculitis Translational Research Section, National Institute of Arthritis and Musculoskeletal and Skin Disease, National Institutes of Health; Columbia University Vagelos College of Physicians and Surgeons; Laboratory of Allergic Diseases, National Institute of Allergy and Infectious Diseases, National Institutes of Health; Phoenix Children’s Hospital; University of Arizona; Immune Deficiency Genetics Section, Laboratory of Clinical Immunology and Microbiology, National Institute of Allergy and Infectious Diseases, National Institutes of Health; Faculty of Medicine, Clinica Alemana Universidad del Desarrollo; Division of Immunology and Rheumatology, Hospital Roberto del Rio; Molecular Development of the Immune System Section, National Institute of Allergy and Infectious Diseases, National Institutes of Health; Division of Clinical Medicine, National Institute of Allergy and Infectious Diseases, National Institutes of Health; Office of the Scientific Director, National Institute of Allergy and Infectious Diseases, National Institutes of Health; Research Technologies Branch, Collaborative Bioinformatics Resource, National Institute of Allergy and Infectious Diseases, National Institutes of Health; Human Immunological Diseases Section, National Institute of Allergy and Infectious Diseases, National Institutes of Health; University of Pittsburgh, Division of Rheumatology and Clinical Immunology

**Author notes:** Corresponding author: Christopher Redmond, National Institute of Arthritis and Musculoskeletal and Skin Diseases National Institutes of Health, 9000 Rockville Pike, Bethesda, MD 20892.

## Abstract

Multisystem Inflammatory Syndrome in Children (MIS-C) is a systemic inflammatory condition that follows SARS-CoV2 infection or exposure in children. Clinical presentations are highly variable and include fever, gastrointestinal (GI) disease, shock, and Kawasaki Disease-like illness (MIS-C/KD). Compared to patients with acute COVID, patients with MIS-C have a distinct immune signature and expansion of *TRVB11* expressing T cells. However, the relationship between immunological and clinical phenotypes of MIS-C is unknown. Here, we measured serum biomarkers, TCR repertoire, and SARS-CoV2-specific T cell responses in a cohort of 76 MIS-C patients. Serum biomarkers associated with macrophage and Th1 activation were elevated in patients with shock, consistent with previous reports. Significantly increased SARS-CoV-2-induced IFN-γ, IL-2, and TNF-α production were seen in CD4^+^ T cells from patients with neurologic involvement and respiratory failure. Diarrhea was associated with a significant reduction in shock-associated serum biomarkers, suggesting a protective effect. *TRVB11* usage was highly associated with MIS-C/KD and coronary aneurysms, suggesting a potential biomarker for these manifestations in MIS-C patients. By identifying novel immunologic associations with the different clinical phenotypes of MIS-C, this study provides insights into the clinical heterogeneity of MIS-C. These unique immunophenotypic associations could provide biomarkers to identify patients at risk for severe complications of MIS-C, including shock and MIS-C/KD.

## Research Letter

Multisystem Inflammatory Syndrome in Children (MIS-C) is a systemic inflammatory condition seen in children following severe acute respiratory syndrome coronavirus 2 (SARS-CoV-2) infection or exposure. Symptoms are highly variable and include fever, diarrhea, cardiogenic shock, respiratory compromise, and Kawasaki Disease (KD)-like features (1, 2). Compared with acute Coronavirus Disease 2019 (COVID-19), MIS-C is characterized by distinct immune responses, including increased monocyte/macrophage-derived cytokines such as interleukin (IL) -6 and IL-18, T-cell-derived cytokines including IL-10 and IL-17A, and *TRβ V11-2* expressing T cells (3, 4). However, the relationship between MIS-C-related immunophenotypes and clinical heterogeneity is unknown.

We determined clinical associations of MIS-C immunophenotypes in a cohort of 76 MIS-C patients (4). Clinical characteristics of the cohort were previously reported (4) and used to identify patients with diarrhea, cardiac, respiratory, neurologic, and KD-like symptoms (**Table S1, S2**). Serum biomarkers (n = 69) and T cell receptor (TCR) repertoire (n = 58) were measured as previously reported (4). To quantify SARS-CoV-2-specific T cell responses (n = 24), we stimulated peripheral blood mononuclear cells (PBMCs) with spike, membrane, or nucleocapsid SARS-CoV-2 peptides; cytokine expression was measured using intracellular staining and flow cytometry. Groups were compared using Mann-Whitney with multiple comparison adjustment.

Serum biomarkers associated with macrophage and Th1 activation, including IFN-γ, were elevated in patients with shock, consistent with previous reports (3-5) (**Fig 1A, S1**). Accordingly, significantly increased SARS-CoV-2-induced IFN-γ, IL-2, and TNF-α production were seen in CD4^+^ T cells from patients with neurologic involvement and respiratory failure (**Fig 1B-C, S1**). SARS-CoV-2-specific IFN-γ production was also seen in CD8^+^ T cells from patients with neurologic involvement (**Fig S2**). No significant antigen-specific responses were seen in CD4^+^-derived IL-10 and IL-17A, or in CD8^+^-derived IL-2, or TNF-α (**Fig S1**). Unexpectedly, diarrhea was associated with a significant reduction in shock-associated serum biomarkers, but not with reduced antigen-specific T cell responses (**Fig 1A, S1**). *TRVB11-2* gene usage was significantly associated with KD and coronary aneurysms (**Fig 1D**).

**Figure 1:**
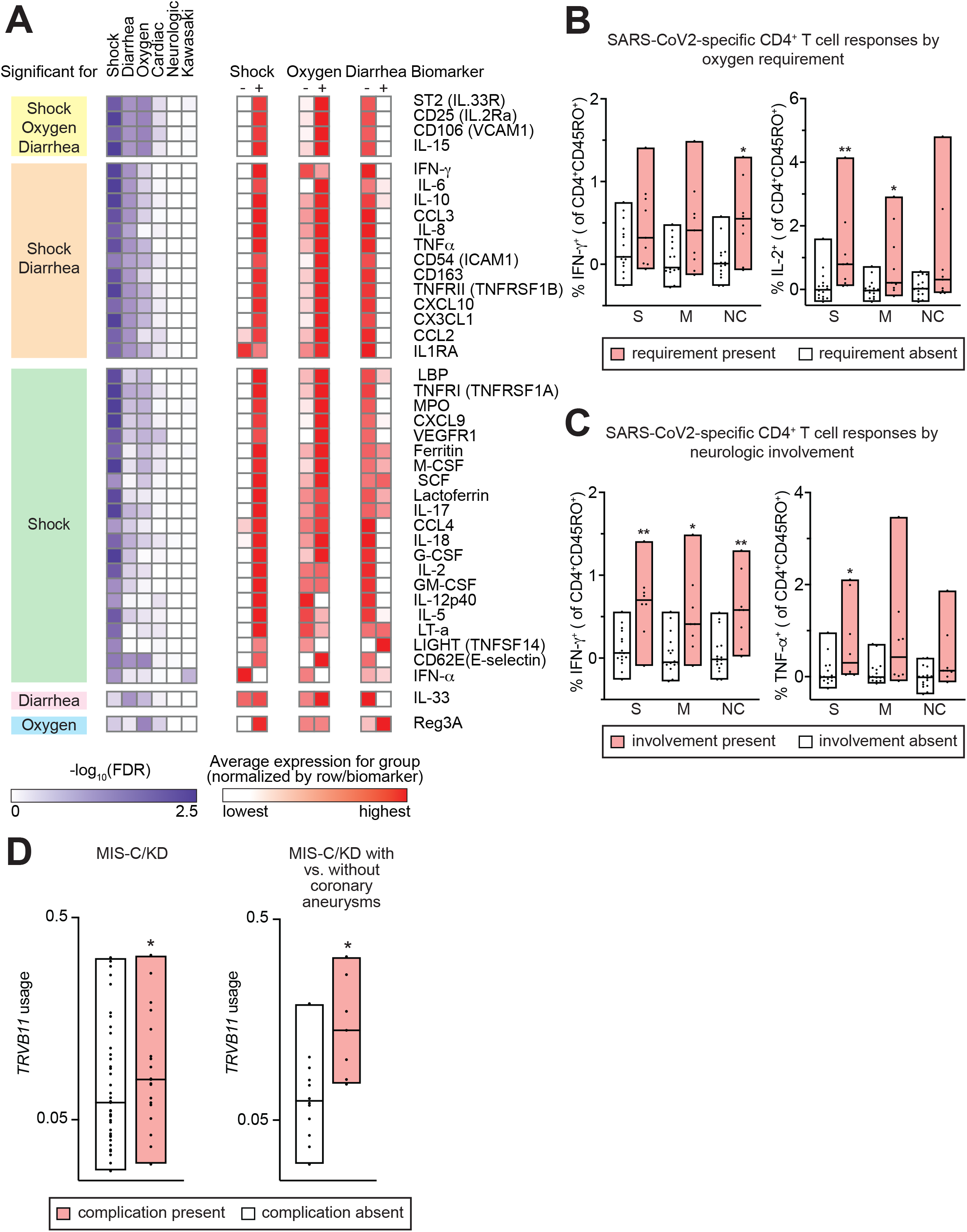
Clinical associations with immunological phenotypes in MIS-C. A. Serum biomarkers significantly associated with clinical phenotypes in MIS-C. Biomarkers are grouped by clinical associations. Heatmap (purple) displays significance of clinical association. Heatmap (red) displays average expression level of each biomarker for the clinical group, normalized by row. B,C. Bar graphs display CD4^+^CD45RO^+^ (memory) T cell responses to SARS-CoV2 peptides in patients with oxygen requirement (B) or neurological involvement (C). PBMC were stimulated for 6h in the presence of CD28/CD49d alone or in combination with spike, membrane, or nucleocapsid peptide pools. Cytokine expression was measured using intracellular staining and flow cytometry, and the difference was calculated between CD28/CD49d + peptide-treated and CD28/CD49d-treated cells. Responses were compared in subjects with vs. without clinical symptoms. D. Bar graphs display *TRVB11* usage in MIS-C patients with vs. without Kawasaki Disease-like phenotype (MIS-C/KD), and in MIS-C/KD patients with vs. without coronary artery aneurysms. *FDR<0.05, **FDR<0.01, Mann-Whitney with multiple comparison adjustment.

By identifying novel associations with different phenotypes, this study gives insights into potential immunologic mechanisms underlying the clinical heterogeneity of MIS-C. The elevation of serum inflammatory markers – particularly IFN-γ – in patients with severe disease is consistent with findings in other cohorts (2-5). SARS-CoV-2-specific CD4^+^ responses in patients with neurological and respiratory involvement suggests that antigen-specific T cell functions may contribute to these symptoms. Conversely, serum biomarker data indicates that diarrhea may protect from severe inflammation in MIS-C, independent of T cell responses. While the GI tract can function as a SARS-CoV-2 antigen reservoir, diarrhea does not always correlate with antigenemia and may portend milder disease in acute COVID-19 (6). Finally, the novel association of MIS-C/KD with expansion of *TRVB11-2* -expressing T cells – which is thought to reflect superantigenic responses to spike glycoprotein (7) – suggests a potential link between superantigens and MIS-C/KD, and a possible biomarker for this severe manifestation.

Limitations of this study include variable timing of sample acquisition relative to hospitalization, with samples drawn significantly earlier from patients with shock and neurologic involvement. However, associations with shock were seen in previous studies (2-5), and other phenotypes were not significantly associated with timing relative to hospitalization. Another potential confounder is that 96% of patients were treated with systemic immunomodulators. Nonetheless, inflammatory responses were still detected because samples were collected during a period of active disease. Strengths include the use of a large multicenter international cohort that improved the generalizability of these results and deep clinical and immunologic profiling that enabled discovery of novel associations. As MIS-C remains a significant complication even in the post-vaccine era (1), further investigations should refine biomarkers for severe manifestations, target immunomodulatory treatment, and improve outcomes.

## Supporting information

Supplementary Data (all)

## Data Availability

All data produced in the present study are available upon reasonable request to the authors

## Acknowledgements

The authors would like to thank the other members of the NIAID COVID-19 consortium, the referring physicians from partner institutions, and the patients for their participation in this study. The authors would like to thank Laura Lewandowski for her thoughtful review and comments.

## Author Declarations

The authors declare no competing interests

## Ethical Approval and Consent

This was a natural history study of patients with MIS-C enrolled at the referring institutions. Informed consent was provided by the parents or guardians and/or assent by minors. Samples size was not predetermined. Blood samples were collected between Mar 2020 and Feb 2021 under protocols approved by the following institutional review boards (IRBs): Comité Ético Científico Facultad de Medicina Clínica Alemana Universidad del Desarrollo, Santiago, Chile (protocol 2020-41); Ethics Committee of the Fondazione IRCCS Policlinico San Matteo, Pavia, Italy (protocol 20200037677); Comitato Etico Interaziendale A.O.U. Città della Salute e della Scienza di Torino, Turin, Italy (protocol 00282/2020); Ethics Committee of the University of Naples Federico II, Naples, Italy (protocol 158/20); Comitato Etico Provinciale, Brescia, Italy (protocol NP-4000); University of Milano Bicocca-San Gerardo Hospital, Monza, and Ethics Committee of the National Institute of Infectious Diseases ‘Lazzaro Spallanzani’, Italy (protocol 84/2020); Hadassah Medical Organization IRB, Jerusalem, Israel (protocol HMO-235-20); and National Institute of Allergy and Infectious Diseases (NIAID), National Institutes of Health (NIH), Bethesda, Maryland, USA (protocols <u>NCT04582903</u>, <u>NCT03394053</u> and <u>NCT03610802</u>). All identifiers included in this study are study identifiers that cannot reveal the identity of the research subjects and are publicly available through other works published by the NIAID COVID consortium (4).

## References

1. Nygaard U, Holm M, Hartling UB, Glenthøj J, Schmidt LS, Nordly SB, et al. Incidence and clinical phenotype of multisystem inflammatory syndrome in children after infection with the SARS-CoV-2 delta variant by vaccination status: a Danish nationwide prospective cohort study. The Lancet Child & Adolescent Health. 2022;6(7):459–65.

2. Consiglio CR, Cotugno N, Sardh F, Pou C, Amodio D, Rodriguez L, et al. The Immunology of Multisystem Inflammatory Syndrome in Children with COVID-19. Cell. 2020;183(4):968–81 e7.

3. Hoste L, Roels L, Naesens L, Bosteels V, Vanhee S, Dupont S, et al. TIM3+ TRBV11-2 T cells and IFNgamma signature in patrolling monocytes and CD16+ NK cells delineate MIS-C. J Exp Med. 2022;219(2).

4. Sacco K, Castagnoli R, Vakkilainen S, Liu C, Delmonte OM, Oguz C, et al. Immunopathological signatures in multisystem inflammatory syndrome in children and pediatric COVID-19. Nat Med. 2022;28(5):1050–62.

5. Diorio C, Shraim R, Vella LA, Giles JR, Baxter AE, Oldridge DA, et al. Proteomic profiling of MIS-C patients indicates heterogeneity relating to interferon gamma dysregulation and vascular endothelial dysfunction. Nat Commun. 2021;12(1):7222.

6. Britton GJ, Chen-Liaw A, Cossarini F, Livanos AE, Spindler MP, Plitt T, et al. Limited intestinal inflammation despite diarrhea, fecal viral RNA and SARS-CoV-2-specific IgA in patients with acute COVID-19. Sci Rep. 2021;11(1):13308.

7. Porritt RA, Paschold L, Rivas MN, Cheng MH, Yonker LM, Chandnani H, et al. HLA class I-associated expansion of TRBV11-2 T cells in multisystem inflammatory syndrome in children. J Clin Invest. 2021;131(10).

