## Supplementary Data (all) for "Deep immune profiling uncovers novel associations with clinical phenotypes of Multisystem Inflammatory Syndrome in Children (MIS-C)"

**Table S1: Kawasaki Disease criteria in MIS-C Cohort**

| Patient ID | Bilateral conjunctival injection | Oral mucous membrane changes | Cervical lymph-adenopathy | Peripheral extremity changes | Poly-morphous rash | Coronary Dilatation | Abnormal supplemental laboratory findings | Final Determination: MISC/KD? |
| --- | --- | --- | --- | --- | --- | --- | --- | --- |
| PIMS-019 | No | No | No | No | No | No | No | No |
| Israel_P5 | No | No | No | No | No | No | No | No |
| PIMS-042 | No | No | No | No | No | No | No | No |
| PIMS-008 | No | No | No | No | No | No | No | No |
| NAP012 | No | No | No | No | No | No | No | No |
| Israel_P4 | No | No | No | No | No | No | No | No |
| Israel_P3 | No | No | No | No | No | No | No | No |
| PIMS-005 | No | No | No | No | No | No | No | No |
| PIMS-024 | No | No | No | No | No | No | No | No |
| Israel_P2 | No | No | No | No | No | No | No | No |
| Israel_P1 | No | No | No | No | No | No | No | No |
| NAP013 | No | No | No | No | No | No | No | No |
| PIMS-014 | NA | No | No | No | No | No | No | No |
| TO-036 | Yes | No | No | No | No | No | No | No |
| TO-051 | Yes | No | No | No | No | No | No | No |
| PIMS-004 | No | No | No | No | Yes | No | No | No |
| TO-039 | Yes | No | No | No | No | No | No | No |
| PIMS-029 | Yes | No | No | No | No | No | No | No |
| PIMS-038 | No | No | No | No | Yes | No | No | No |
| PIMS-022 | NA | No | NA | No | Yes | NA | No | No |
| PIMS-003 | No | No | No | No | Yes | No | No | No |
| PIMS-039 | Yes | No | No | No | No | No | No | No |
| TO-043 | Yes | No | No | No | No | No | No | No |
| TO-017 | Yes | No | No | No | No | No | No | No |
| OD0079 | Yes | No | No | No | No | No | No | No |
| TO-034 | Yes | No | No | No | No | No | No | No |
| PIMS-044 | No | No | No | No | Yes | No | No | No |
| PIMS-031 | No | Yes | No | No | Yes | No | No | No |
| PIMS-055 | Yes | No | No | No | Yes | No | No | No |
| PIMS-013 | Yes | No | No | No | Yes | No | No | No |
| TO-024 | Yes | No | No | No | Yes | No | No | No |
| PIMS-001 | No | Yes | No | No | Yes | No | No | No |
| TO-027 | Yes | No | No | No | Yes | No | No | No |
| PIMS-030 | Yes | No | No | No | Yes | No | No | No |
| PIMS-036 | No | Yes | No | No | Yes | No | No | No |
| PIMS-007 | Yes | No | No | No | Yes | No | No | No |
| PIMS-016 | Yes | No | No | No | Yes | No | No | No |
| PIMS-006 | Yes | No | No | No | Yes | No | No | No |
| TO-052 | Yes | No | No | No | Yes | No | No | No |
| OD0086 | Yes | No | No | No | Yes | No | No | No |
| TO-040 | No | Yes | Yes | Yes | No | No | No | No |
| TO-054 | Yes | No | Yes | No | Yes | No | No | No |
| TO-045 | Yes | No | Yes | No | Yes | No | No | No |
| PIMS-049 | Yes | Yes | No | No | Yes | No | No | No |
| TO-001 | Yes | Yes | Yes | No | No | No | No | No |
| TO-041 | Yes | No | Yes | No | Yes | No | No | No |
| TO-010 | Yes | Yes | No | No | Yes | No | No | No |
| PIMS-050 | Yes | Yes | No | No | Yes | No | No | No |
| TO-042 | Yes | Yes | Yes | No | No | No | Yes | Yes |
| TO-055 | Yes | Yes | Yes | No | Yes | No | No | Yes |
| PIMS-010 | Yes | Yes | No | Yes | Yes | No | No | Yes |
| TO-049 | Yes | Yes | Yes | No | Yes | No | No | Yes |
| TO-056 | Yes | Yes | No | Yes | Yes | No | No | Yes |
| TO-053 | Yes | Yes | No | Yes | Yes | No | No | Yes |
| PIMS-017 | Yes | Yes | No | No | Yes | No | Yes | Yes |
| TO-032 | Yes | Yes | Yes | No | Yes | No | No | Yes |
| TO-048 | Yes | Yes | No | Yes | Yes | No | No | Yes |
| TO-006 | Yes | Yes | Yes | No | Yes | No | No | Yes |
| PIMS-046 | Yes | Yes | No | No | Yes | No | Yes | Yes |
| TO-044 | Yes | Yes | Yes | Yes | No | No | No | Yes |
| TO-035 | Yes | Yes | No | No | Yes | No | Yes | Yes |
| OD0092 | No | Yes | Yes | No | Yes | No | Yes | Yes |
| NAP044 | No | No | No | No | No | Yes | No | Yes |
| TO-037 | Yes | No | No | Yes | Yes | No | Yes | Yes |
| Israel_P6 | Yes | Yes | No | Yes | Yes | No | No | Yes |
| TO-009 | Yes | Yes | Yes | Yes | Yes | No | No | Yes |
| OD0077 | Yes | No | No | No | No | Yes | No | Yes |
| PV-006 | No | Yes | No | No | Yes | Yes | No | Yes |
| PIMS-027 | Yes | No | No | No | Yes | Yes | No | Yes |
| PIMS-023 | Yes | No | No | No | Yes | Yes | No | Yes |
| PIMS-028 | Yes | No | No | No | Yes | Yes | No | Yes |
| NAP014 | No | Yes | No | Yes | No | Yes | No | Yes |
| TO-046 | Yes | No | No | No | Yes | Yes | No | Yes |
| PV-009 | Yes | No | No | Yes | Yes | Yes | No | Yes |
| PIMS-032 | Yes | Yes | No | No | Yes | Yes | No | Yes |
| NIHPID0111 | No | Yes | No | Yes | Yes | Yes | No | Yes |

| Clinical Phenotype | Classification Criteria |
| --- | --- |
| Respiratory | Any documented use of supplemental oxygen, ranging from nasal cannula oxygen to mechanical ventilation |
| Neurologic | Any documented neurologic symptoms, ranging from headache to encephalopathy |
| Cardiac | Documented myocarditis or cardiac dysfunction |
| Diarrhea | Documentation of diarrhea |
| MIS-C/KD | Patients with documented symptoms and laboratory values meeting the AHA criteria for complete or incomplete KD |
| Shock | Documentation of shock requiring in intensive volume resuscitation or vasopressors |

**Table S2.** **Classification criteria for clinical symptoms**. Classification of clinical symptoms was based on documentation of patients meeting the above criteria. MIS-C/KD classification was based on the 2017 American Heart Association (AHA) criteria for complete and incomplete Kawasaki Disease.

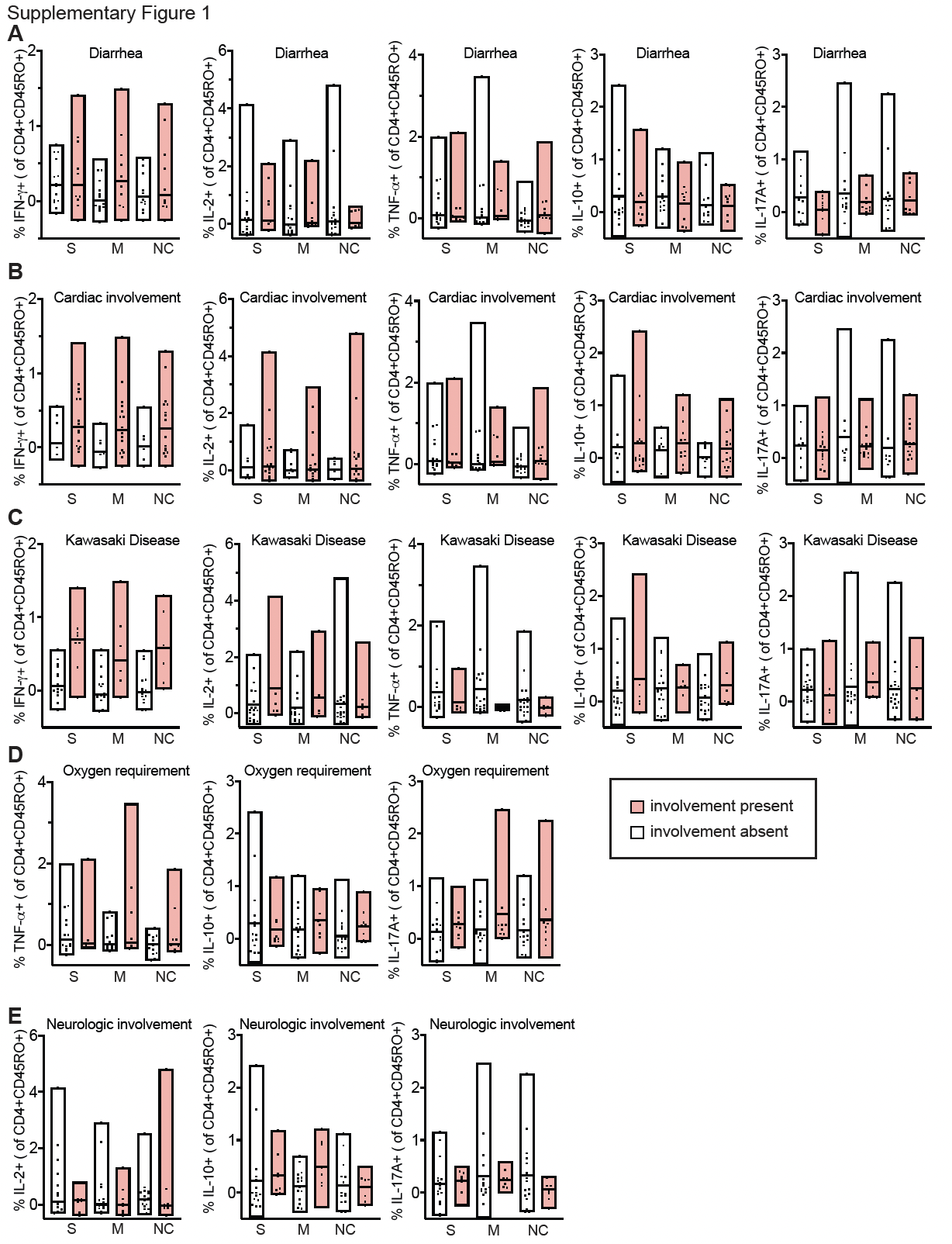

**Figure S1: Clinical associations with immunological phenotypes in MIS-C.** Bar graphs display CD4^+^CD45RO^+^ (memory) T cell responses to SARS-CoV2 peptides in patients with vs. without diarrhea (A), cardiac involvement (B), Kawasaki Disease (C), Oxygen requirement (D), or neurological involvement (E). PBMC were stimulated for 6h in the presence of CD28/CD49d (fastimmune) alone or in combination with spike (s), membrane (m), or nucleocapsid (nc) peptide pools (Miltenyi). Cytokine expression was measured using intracellular staining and flow cytometry, and the difference was calculated between CD28/CD49d + peptide-treated and CD28/CD49d-treated cells. Responses in CD4+ T cells were compared in subjects with vs. without clinical symptoms. Data for IFN-γ and IL-2 production in patients with oxygen requirement and for IFN-γ and TNF-α production in patients with neurologic involvement can be found in the main figure.

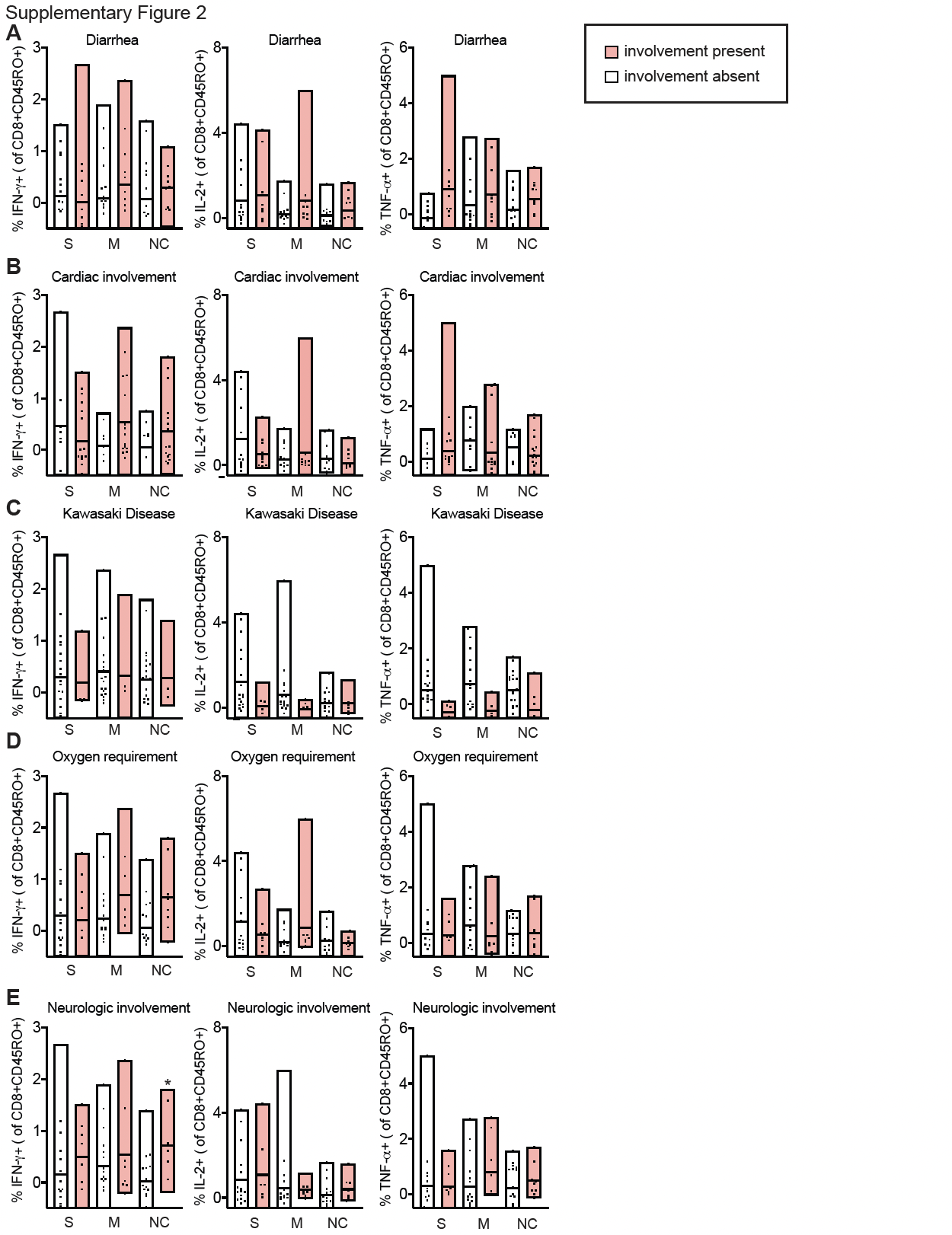

**Figure S2: Clinical associations with immunological phenotypes in MIS-C.** Bar graphs display CD8^+^CD45RO^+^ (memory) T cell responses to SARS-CoV2 peptides in patients with diarrhea (A), cardiac involvement (B), Kawasaki Disease (C), Oxygen requirement (D), or neurological involvement (E). PBMC were stimulated for 6h in the presence of CD28/CD49d (fastimmune) alone or in combination with spike(s), membrane (m), or nucleocapsid (nc) peptide pools (Miltenyi). Cytokine expression was measured using intracellular staining and flow cytometry, and the difference was calculated between CD28/CD49d + peptide-treated and CD28/CD49d-treated cells. Responses in CD8+ T cells were compared in subjects with vs. without clinical symptoms. *FDR<0.05, Mann-Whitney with multiple comparison adjustment.
